# Stability and Motif Analysis of RNA-Seq Reads from COVID-19 Patients

**DOI:** 10.1101/2025.10.16.25338102

**Authors:** Adrian Lee, Emanuele Raggi, Alger M. Fredericks, Alfred Ayala, Jaewook Shin, Gerard J. Nau, Maya Cohen, Mitchell M. Levy, William G. Fairbrother, Sean F. Monaghan

## Abstract

RNA Sequencing (RNA-Seq) can facilitate prompt and precise management of many illnesses, including infection, especially when coupled with PCR. To further optimize RNA-Seq for the purposes of creating a RNA-Seq-informed PCR test, identifying reliable RNA primer targets is paramount. Essential criteria for constructing RNA primer targets include high gene expression or stability of transcripts. We hypothesize that free energy evaluation and motif analysis can demonstrate gene expression level or stability of transcripts to construct ideal RNA primers for RNA-Seq-informed PCR that can expedite the diagnosis and treatment of infection. Whole blood samples were collected from patients diagnosed with COVID-19 in the ICU at Rhode Island Hospital, stored in Paxgene tubes to preserve the integrity of the specimens, and submitted for RNA-Seq by a commercial sequencing service (Azenta/Genewiz). After quality assurance and quality control (QA/QC) measures are performed, we used RNAfold from the ViennaRNA Package to calculate the minimum free energy (MFE) and ensemble free energy (EFE) values to compare the stability among different secondary structures constructed from RNA-Seq read sequences. Energy parameters for calculations were set at 37 degree Celsius and standard physiological conditions at pH 7.4. In our analysis, we utilized multiple statistical tools, such as analysis of variance, Games-Howell post hoc tests, and a negative binomial regression. Of the 676 reads that aligned with SARS-CoV-2, there were 137 unique sequences among all patients. Among the unique read sequences, the average MFE was -30.46 kcal/mol and the average EFE was -32.94 in kcal/mol. There were 6 genes with these unique read sequences, two of which -- nucleocapsid (*N*) gene and spike (*S*) gene -- coded for integral structural proteins. Notably, the mean MFE and EFE of *N* gene were significantly different from *ORF1ab* gene (p = 2.81e-7; p = 0.005) and *ORF6* gene (p = 0.023; p = 0.03). Our motif analysis demonstrated 7 motifs that corresponded to destabilization of the RNA, and a single motif (MEME-28) that corresponded to stabilization of the RNA. Within the nucleocapsid (*N*) gene, we found that reads from different regions also differed in stability. Our results demonstrate that the stability of reads from different genes varies in SARS-CoV-2 infections. Free energy evaluation and motif analysis are viable approaches to determine RNA structure stability or levels of differential gene expression. Stable transcripts that are highly expressed can be ideal RNA primers to be used in conjunction with PCR to expedite the management of infection.

## 1 Introduction

RNA Sequencing (RNA-seq) has introduced an unprecedented realm of research with its in-depth view of the transcriptome. It not only identifies alterations in expressed parts of the genome, but also detects transcript isoforms from alternative splicing, chimeric gene fusions, and novel RNA transcripts (Byron et al., 2016; Mortazavi et al., 2008). In addition, certain regulatory RNAs have been shown to modulate metabolic and virulence functions of pathogens, demonstrating the promising application of RNA-Seq to clinical realms (Oliva et al., 2015; Papenfort et al., 2014). Thus, RNA-Seq has been increasingly incorporated in clinical diagnoses and management (Jonkhout et al., 2021; Peymani et al., 2022; Zhou et al., 2023). Main examples include cancer and infectious diseases.

In breast cancer, RNA-Seq can be used for prognostication via multigene mRNA-based assays and for identifying alternative breast cancer 1 (*BRCA1*) transcripts that were not apparent by conventional methods. In prostate cancer and glioblastoma, RNA-Seq helped detect alternative transcripts that can have therapeutic implications. In chronic myeloid leukemia (CML), RNA-Seq helped identify previously undetected resistance mutations that can influence treatment (Byron et al., 2016).

In the realm of infectious diseases, RNA-Seq has contributed to detailed profiling of immune cells in bacterial, viral, parasitic, and fungal infections. For instance, profiling of infected macrophages revealed that gene expression heterogeneity creates multiple environments for Salmonella to survive and exploit the host. RNA-Seq also allowed high-resolution mapping of Plasmodium spp. life cycle, identifying potential targets for vaccine and anti-parasitic therapies. Though still in infancy, research in Candida showed a possible candidemia-risk allele which may discourage the migration of monocytes (Huang et al., 2021).

Polymerase Chain Reaction (PCR) tests have been used to amplify and identify the DNA of pathogens in targeted sites such as the respiratory tract (Camargo et al., 2019; Covert et al., 2021; Palavecino et al., 2020). RNA-based molecular tests have the potential to expedite diagnoses and treatments since, compared to DNA, RNA is more abundant, can be a real-time marker for active infection, and represent expressed genes and isoforms which are readily identified with RNA-Seq. Especially in sepsis care where mortality rate rises exponentially every hour, RNA-Seq-informed PCR tests have the potential to identify pathogens more efficiently and precisely than blood cultures, which take 3-5 days.

Our lab previously demonstrated that RNA-Seq can establish the presence of severe acute respiratory syndrome coronavirus 2 (SARS-CoV-2) and define the host’s response in COVID-19 patients, thereby identifying genes, proteins, and entropy as potential biomarkers for diagnosis and therapeutics. Two most abundant genes in blood found were the RNA Dependent RNA Polymerase and the N protein, with the latter showing the highest number of reads (Fredericks et al., 2022). It remains unclear what factors determine the extent of repeated sequences in an experiment, especially with the numerous processing steps necessary in the RNA-Seq procedure (Fu et al., 2018) though it is common that read sequences may be repeatedly identical (Deschamps-Francoeur et al., 2020). The reason for such abundance can be surmised as either baseline high gene expression or stability of its transcripts, both of which can contribute to constructing a reliable RNA primer for the RNA-Seq-informed PCR (Fredericks et al., 2022).

In this study, we utilized transcriptome data collected from COVID-19 patients in the ICU using deep RNA-Seq to identify differences of read structure and stability. The COVID-19 genome is relatively small at approximately 30 kb and there are two large overlapping open reading frames that encode a total of 16 nonstructural protein as well as four open reading frames that encode structural proteins. The small size of the genome and few number of gene regions makes for a good size for our focused analysis on the small COVID-19 genome (Wu et al., 2023). The analysis consisted of assessing the stability, which we refer in this publication as the energy level in kcal/mol of the structure of an RNA molecule, measured by the minimum free energy (MFE) and the ensemble free energy (EFE) of the structure (Ding et al., 2005; Doshi et al., 2004; Lorenz et al., 2011; Trotta et al., 2014; Wuchty et al., 1999) and the global entropy of RNA fragments from differentially expressed transcripts in COVID-19 patients. We hypothesize that free energy evaluation and motif analysis can demonstrate high gene expression level or increased stability of transcripts which indicate preferred RNA primers for RNA-Seq-informed PCR that can expedite the diagnosis and treatment of infection.

## 2 Materials and methods

During 2020, whole blood samples were collected from patients in the ICU, stored in Paxgene tubes to preserve the integrity of the specimens, and submitted for RNA sequencing (Fredericks et al., 2022) by a commercial sequencing service (Azenta/Genewiz). Sequencing was performed on Illumina HiSeq machines, providing 150 base pairs, paired end reads. Libraries were prepared by Genewiz using both the Globin Zero Gold (Illumina) and NEBNext Ultra RNA Library Prep Kit (New England Biolabs).

Three samples were used per lane, and each lane produced 250 million reads. Each sample had >100 million reads per lane. There were 15 total patients, 7 deceased. After QA/QC measures were performed, we then used RNAfold from the ViennaRNA Package, which utilizes a loop-based energy model and dynamic program algorithm to calculate the minimum free energy (MFE) and ensemble free energy values (EFE) to compare the stability among different secondary structures constructed from RNA-Seq read sequences. MFE can be estimated using the algorithm that produces a single optimal structure and EFE is estimated based on a partition function algorithm (Lorenz et al., 2011; McCaskill, 1990; Zuker et al., 1981).

Energy parameters for calculations were set at 37°C and standard physiological conditions with a pH of 7.4. Statistical analysis was computed using R. For sequences that were outliers in length were disregarded for analysis. Only sequences that were less than 175 nucleotides long were analyzed. This was both due to computational limitations for calculations with longer sequences and that the RNA-Seq kit was only designed for finding short RNA reads. A distribution of length of the reads can be seen in Figure 1.

**Figure 1.**
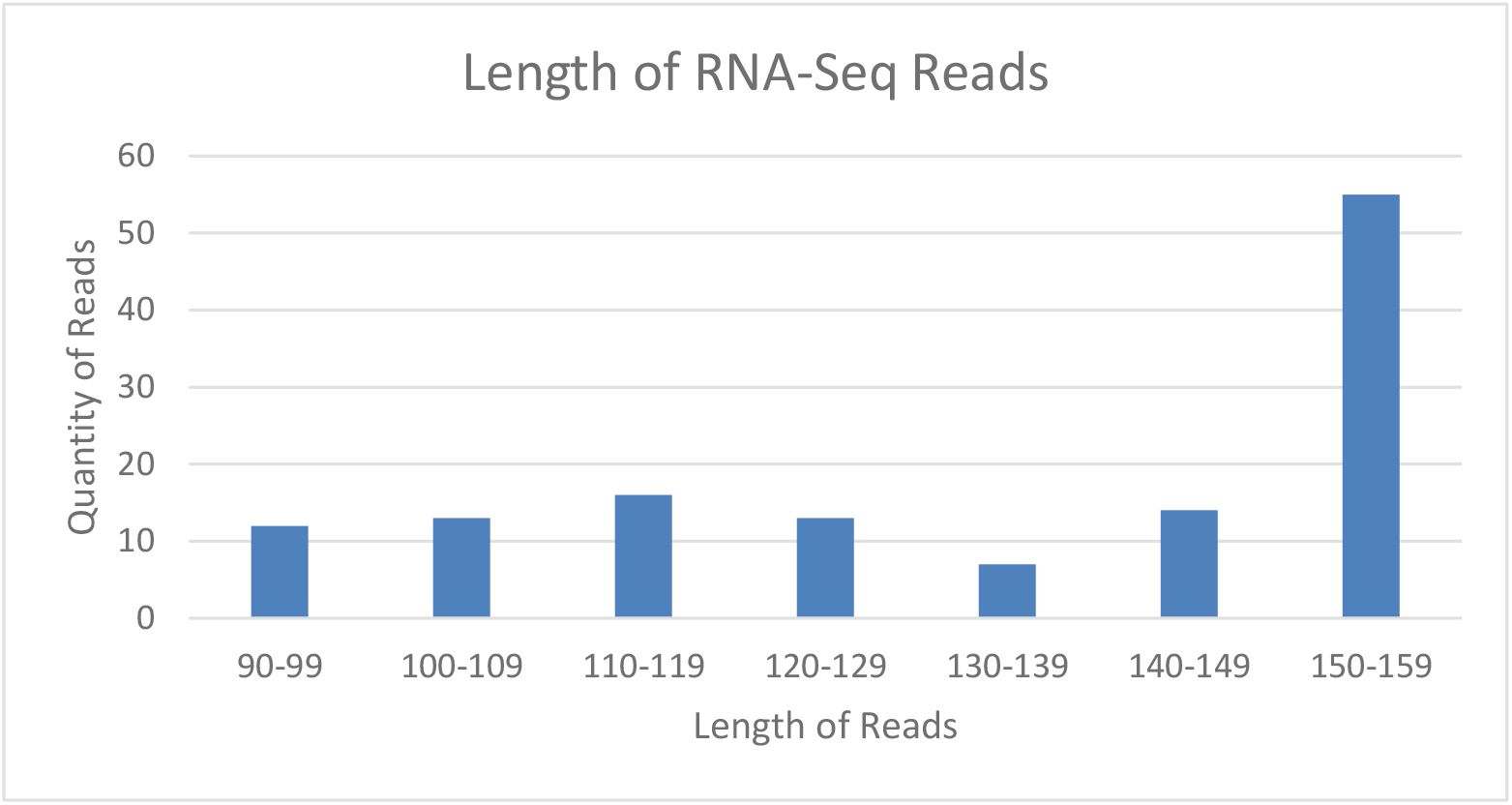
Distribution of the length of reads from the RNA-Seq experiment. Outlier reads longer than 175 nucleotides were omitted from the analysis.

To establish overall differences between read stability from different genes, a Welch ANOVA was first performed to compare MFE and EFE values for reads from different genes. A Games-Howell post hoc test was performed for pairwise comparison of free energy values between different genes. Similarly for a more focused analysis on the *N* gene, a Welch ANOVA was performed to compare MFE values and the EFE values for reads at the beginning, middle, or end of the *N* gene.

The *N* gene was divided into three equally long regions, and allocation to the region of the N gene was determined by the middle of the read sequence. A Games-Howell post hoc test was performed for pairwise comparison of free energy values between regions of the *N* gene. Because the early gene region for the *N* gene had zero variance, a Welch T-test was conducted to compare individual within gene regions. A Bonferroni correction adjustment was utilized due to multiple comparisons between the within gene regions. Separately, a Chi-Squared goodness of fit test was performed to assess the distribution of reads among the genes overall. This established if certain genes had more or fewer reads relative to other genes.

For motif analysis of RNA-Seq read sequences, MEME-ChIP (Machanick et al., 2011) was used to find and analyze motifs and compare with the RNA database. Sequences were entered to the online version 5.5.1 MEME Suite (Bailey et al., 2015). Sequences inputted into MEME-ChIP are ideally 500 letters in length while the longest read sequences used in our analysis was 151 nucleotides long. MEME-ChIP was used to find only the first 10 motifs. Additionally, a newer motif analysis tool called XSTREME (Grant et al., 2021) was used since it allows for input sequences to be of any length. XSTREME utilizes two well established motif discovery programs, MEME and STREME, to identify motifs and utilizes the SEA algorithm for motif enrichment analysis. Sequences were entered to the online version 5.5.1 MEME Suite.

A Welch T-test was performed to compare the MFE and EFE of the sequences with and without the motif. No multiple comparison tests were necessary since these tests were direct comparisons of reads with the motif and without the motif. Analysis of the effect of a destabilizing motif on quantity of duplicated reads was conducted using a negative binomial regression. A Poisson regression was considered for analysis, but the data was over dispersed.

## 3 Results

Of the 676 reads from RNA-Seq, there were 137 unique sequences. In other words, 539 reads were identical with another read. Among all the unique read sequences, the average minimum free energy (MFE) in kcal/mol was -30.46 and the average ensemble free energy (EFE) in kcal/mol was - 32.94. Of the repeated sequences, there was a sequence that was repeated 328 times. The MFE for this sequence was -33.00 kcal/mol and the EFE was -35.20 kcal/mol. Despite being highly repetitive, it was only the 48^th^ lowest MFE and the 60^th^ lowest EFE.

Our Chi-Squared goodness of fit test demonstrated that the proportion of reads from different genes was significantly not equal (p < 2.2e-16), meaning that the genes that were captured by RNA-Seq were captured unevenly. The number of reads per gene and their percentages can be seen in Table 1.

**Table 1.** Table of captured genes, the number of reads per gene, and the percentage of reads from the RNA-Seq experiment per gene.

| Gene Name | Number of Reads | Percentage of Overall Reads |
| --- | --- | --- |
| *ORF1ab* | 143 | 21.2% |
| *ORF3a* | 3 | 0.4% |
| *ORF6* | 10 | 1.5% |
| *ORF8* | 5 | 0.7% |
| *S* | 12 | 1.8% |
| *N* | 454 | 67.2% |
| (Outside of Genes) | 40 | 5.9% |

For the analysis of MFE and EFE values of read sequences in whole gene regions, unique read sequences were found in only six genes: the nucleocapsid (*N*) gene, the *ORF1ab* gene, the *ORF3a* gene, *ORF6* gene, *ORF8* gene, and the spike (*S*) gene. The *N* gene and *S* gene encode integral structural proteins and the *ORF3a, ORF6*, and *ORF8* genes encode auxiliary genes. The *ORF1ab* genes encode other nonstructural proteins. For MFE, A Welch’s ANOVA analysis demonstrated that there was at least one mean MFE for a gene that was significantly different from another gene’s mean MFE (p = 0.0004907). The post hoc analysis assessed fifteen pairs among the six genes and demonstrated three significant relationships. The mean MFE of the *N* gene was significantly different from the mean MFE of the *ORF1ab* gene (p = 2.81e-7) and the *ORF6* gene (p = 0.023). The *ORF6* gene’s mean MFE was significantly different from the *S* gene’s mean MFE (p = 0.037) (Figure 2, 3).

**Figure 2.**
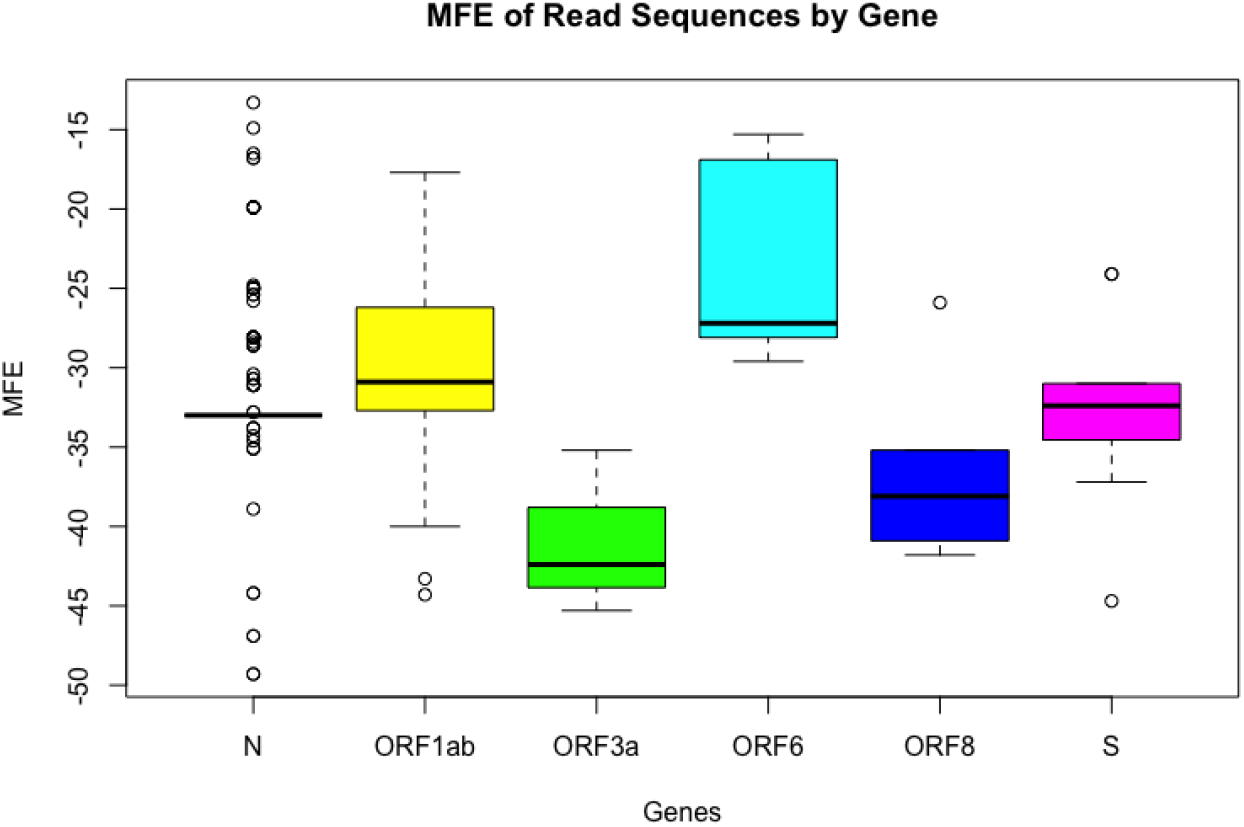
Significant differences of the MFE of RNA-Seq reads in different genes. Due to the highly repeated *N* gene read, the scale of the *N* gene boxplot is shorter with several outliers.

**Figure 3.**
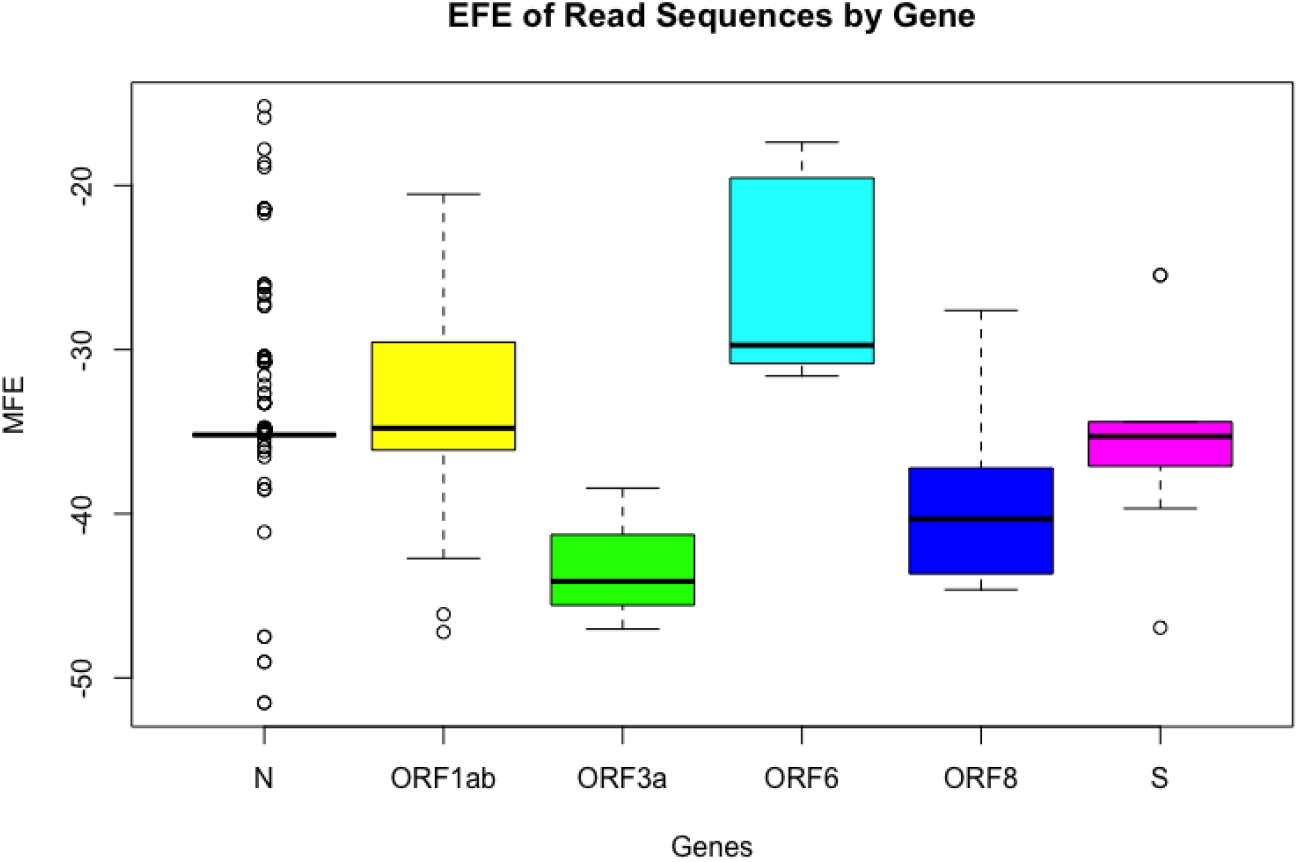
Significant differences of the EFE of RNA-Seq reads in different genes. Due to the highly repeated *N* gene read, the scale of the *N* gene boxplot is shorter with several outliers.

For EFE, our Welch’s ANOVA analysis demonstrated that there was at least one mean EFE for a gene that was significantly different from another gene’s mean EFE (p = 0.002398). The post hoc analysis found four significant pairwise comparisons. The mean EFE of the *N* gene was significantly different from the mean EFE of the *ORF1ab* gene (p = 0.005) and the *ORF6* gene (p = 0.03). The mean EFE of the *ORF3a* gene was significantly different from that of the *ORF6* gene (p = 0.027). The *ORF6* gene’s mean MFE was significantly different from the *S* gene’s mean EFE (p = 0.036). For the analysis of the MFE and EFE values within the *N* gene, a Welch’s ANOVA analysis was initially completed. Since there were only two reads from the early *N* gene region and they were identical, the variance was zero, producing an error in the analysis. To account for this, a Welch’s t-test was completed comparing each of the gene groups individually to each other. A Bonferroni correction adjustment was utilized, and the end region was significantly different from both the early and middle regions in both MFE and EFE values.

The motif analysis using MEME-ChIP of all the read sequences discovered ten motifs, of which six had known or similar motifs in the database (Figure 4, 5). Three of these motifs were found to have a higher MFE for the read sequences (p = 0.00502, p = 0.00000422, p = 0.00023) and a higher EFE for the read sequences (p = 0.0034, p = 0.0000034, p = 0.00039).

**Figure 4.**
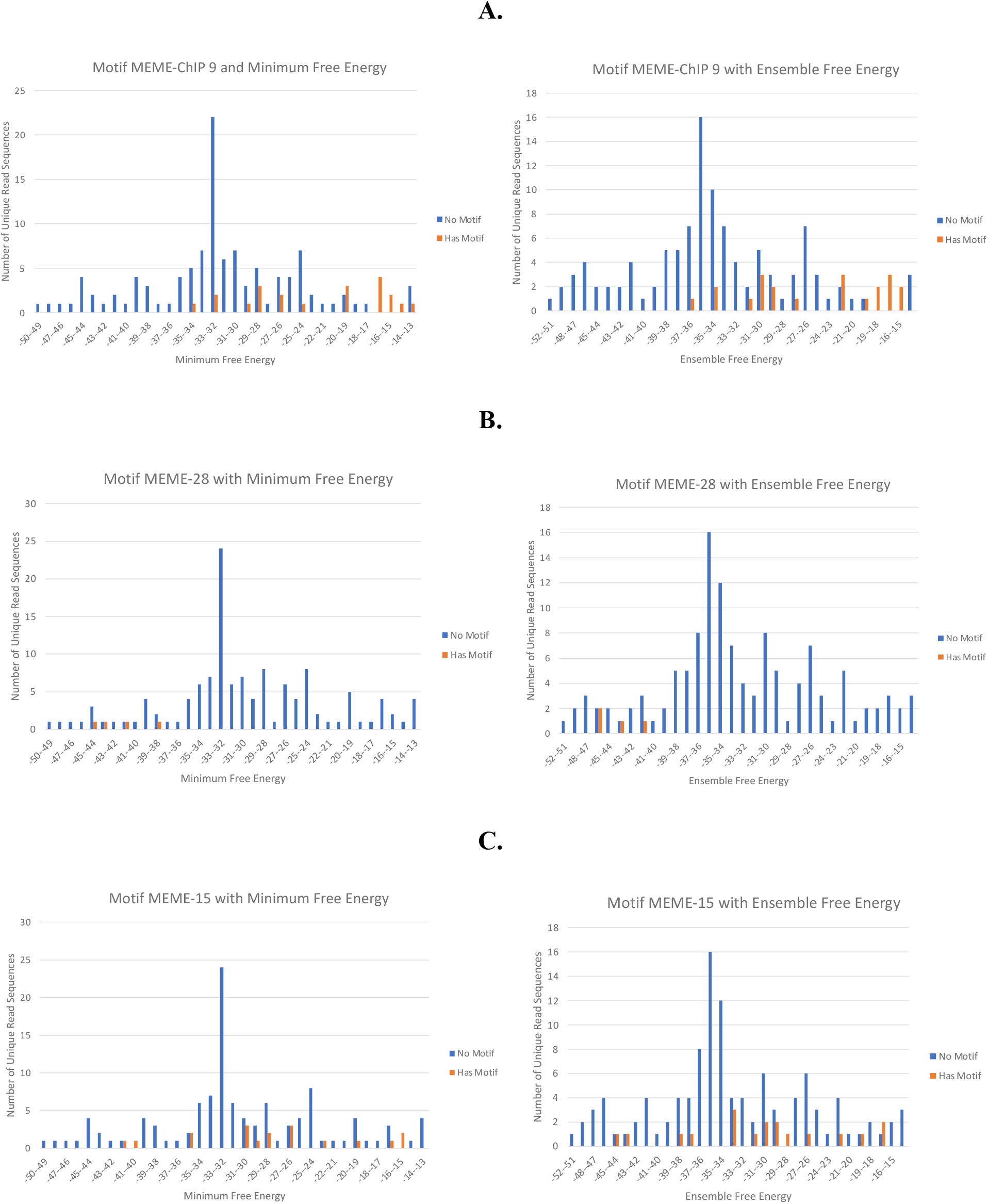
Examples of motif effects on energy. **A.** The motif which we discovered and labeled MEME-ChIP 9 may destabilize RNA sequence reads. (Right) This motif may increase the minimum free energy (p = 0.00000422). (Left) This motif may increase the ensemble free energy (p = 0.0000034). **B**. The motif which we discovered and labeled MEME-28 may stabilize RNA sequence reads. (Right) This motif may decrease the minimum free energy (p = 3.26e-79). (Left) This motif may decrease the ensemble free energy (p = 0.000328). **C**. The motif which we discovered and labeled MEME-15 may not have an effect on RNA sequence read stability. (Right) This motif does not seem to affect the minimum free energy (p = 0.138). (Left) This motif does not seem to affect the ensemble free energy (p = 0.16).

**Figure 5.**
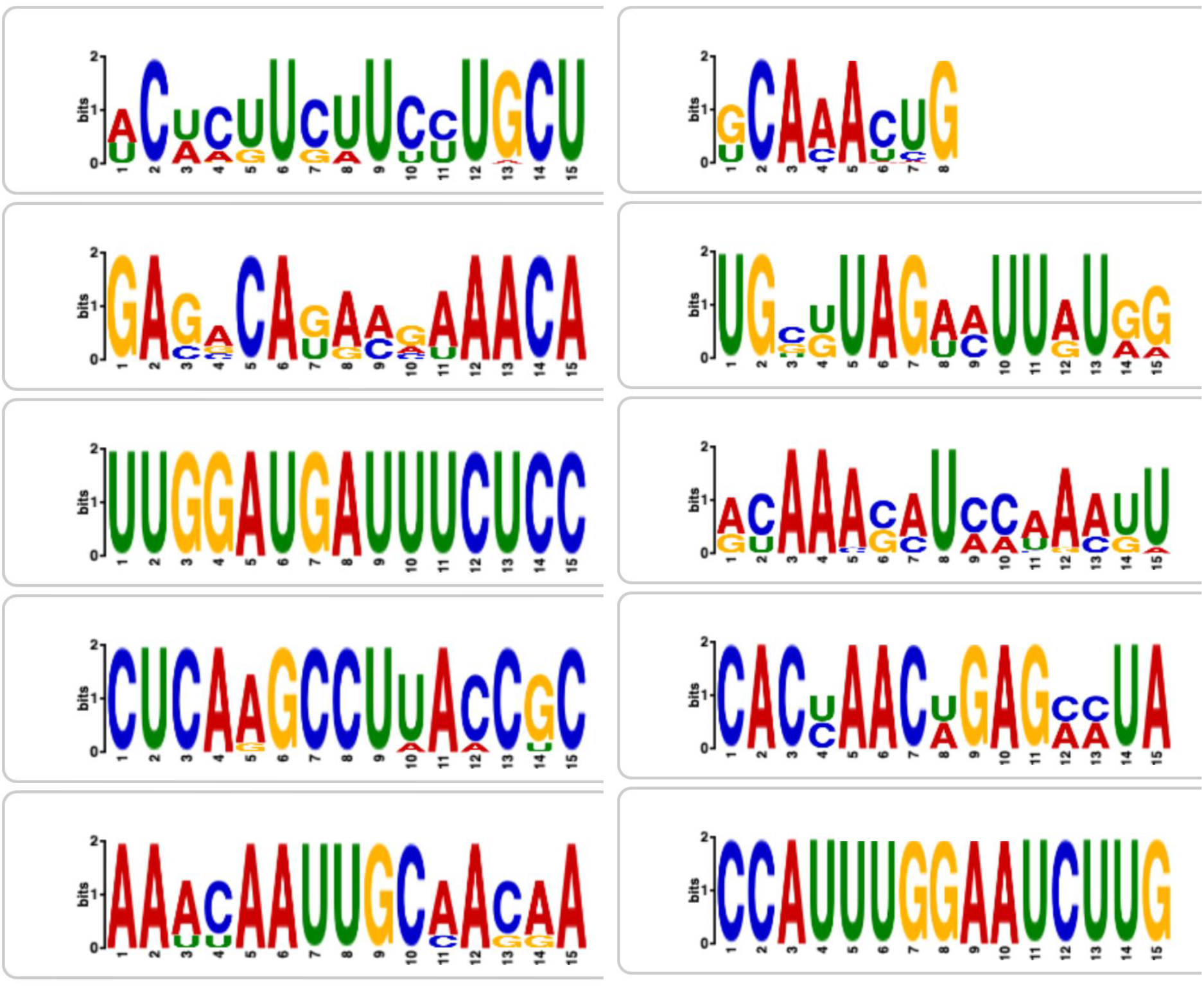
Results from MEME-ChIP analysis. Only ten results were requested to be given with three of them significantly affecting the minimum free energy and ensemble free energy of read sequences.

The motif analysis using XSTREME of all the read sequences discovered 34 motifs (Figure 6). For six of the identified motifs, sequences that had the motifs were found to have significantly different MFE values compared to sequences without the motif (p = 0.0275, p = 0.0204, p = 0.000455, p = 0.0082, p = 0.00175, p = 3.26e-79). These motifs were labeled MEME-9, MEME-10, MEME-21, MEME-22, MEME-27, and MEME-28. For all these motifs besides MEME-10, sequences that had the motifs were also found to have significantly different EFE values compared to sequences without the motif (p = 0.0315, p = 0.000664, p = 0.00695, p = 0.00114, p = 0.000328). These motifs are shown in Table 2 with their IUPAC codes, and because our RNA-Seq reads were consistent across all of the patients in the study, these motifs are also consistent across patients of this study.

**Table 2.**
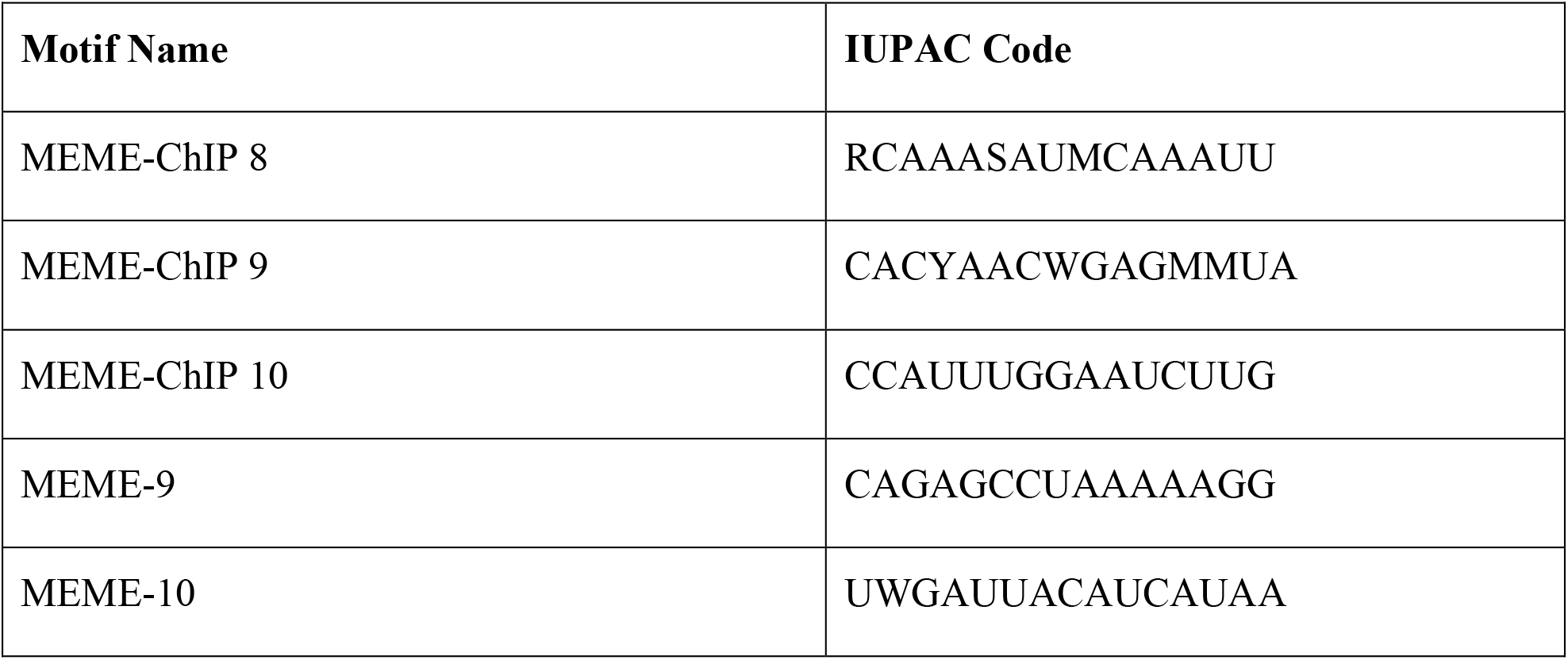
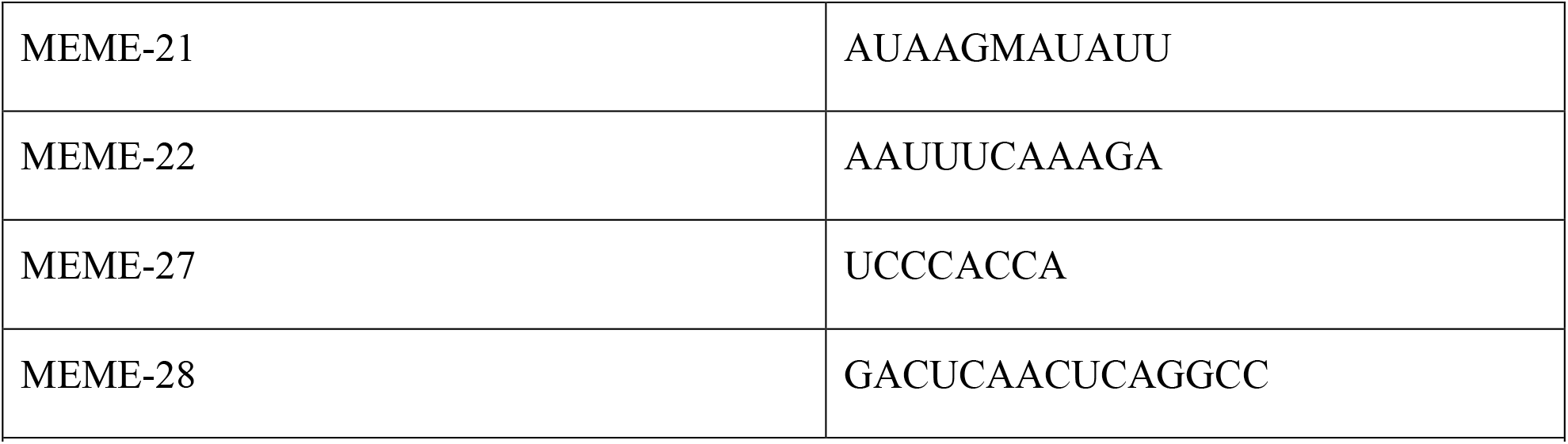
Table of discovered RNA motifs and their IUPAC codes.

**Figure 6.**
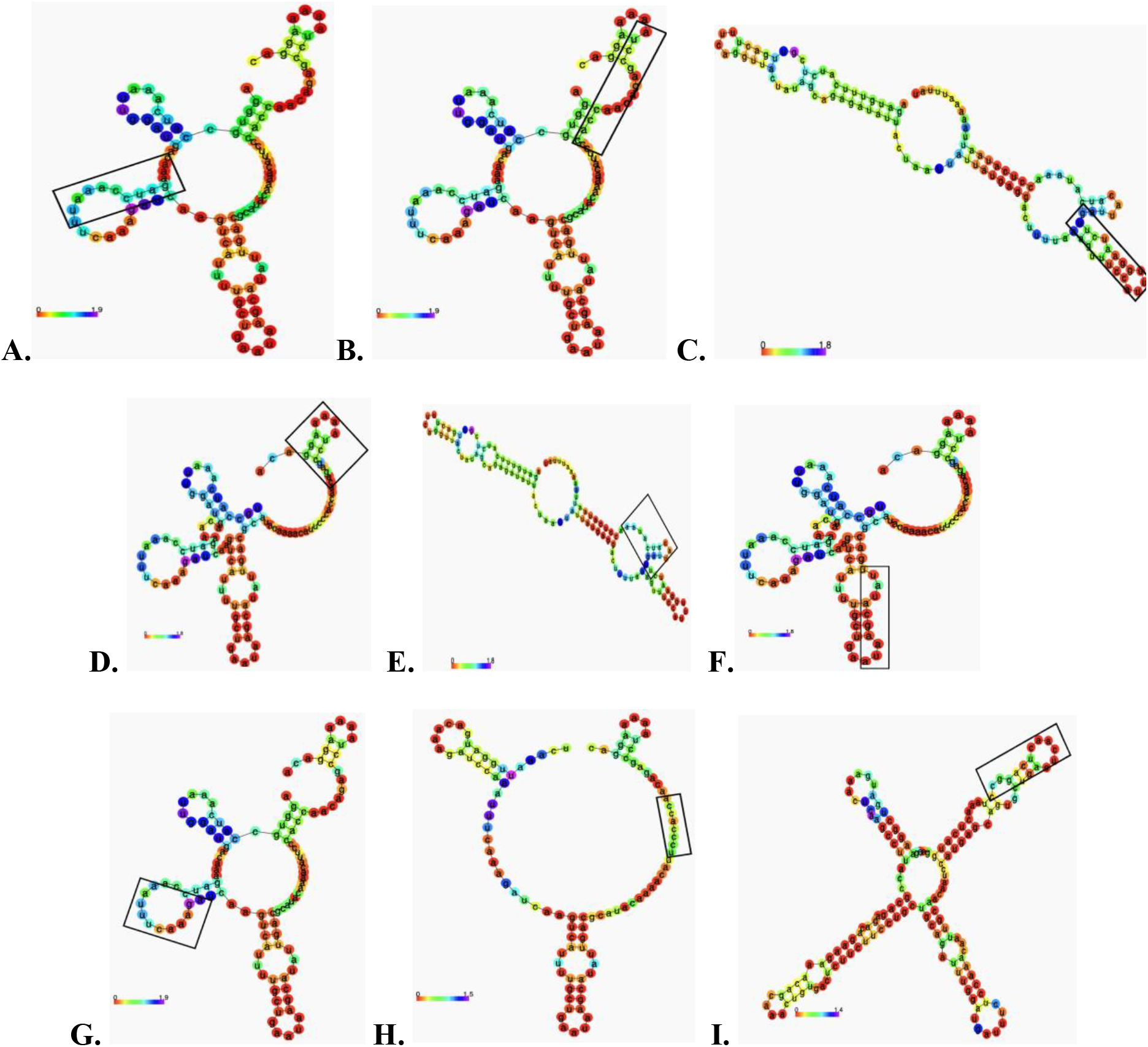
Example secondary structures of reads based on minimum free energy with discovered motifs. **A.** MEME-ChIP 8. **B**. MEME-ChIP 9. **C**. MEME-ChIP 10. **D**. MEME-9. **E**. MEME-10. **F**. MEME-21. **G**. MEME-22. **H**. MEME-27. **I**. MEME-28.

A negative binomial regression was performed to assess if MEME-28, the only motif found to have lower MFE and EFE corresponding to stabilization, was associated with sequences that had a higher number of duplicates. The negative binomial regression formula that was used was: *μμii* =exp(ln(*ttii*)+*ββ*1*xx*1*ii* +*ββ*2*xx*2*ii* +…+*ββkkxxkkii*). This analysis was not significant with a p value of 0.266. A Poisson regression was also considered but the dispersion parameter was calculated to be 161.08, demonstrating overdispersion that negates the statistical significance of the Poisson regression analysis.

## 4 Discussion

To our knowledge, it is unclear if either the stability or high gene expression level accounts for the abundance of RNA-Seq reads. Our chi-squared goodness of fit test demonstrated that the proportion of reads from different genes were not equal, indicating that RNA-Seq does not uniformly collect reads from each gene and that there may be factors that influence a particular sequence being detected from an experiment. Our results indicate that stability of transcripts may play a role in increased detection by RNA-Seq, thus leading to high RNA-Seq reads. It is also possible that with differentially high gene expression may account for the abundance of certain RNA sequences. Thus, if PCR primers were to be designed based on RNA-Seq data, stability of structure should be included not just in design but also for the optimization of the workflow.

RNA-Seq-informed PCR is promising as previous studies have demonstrated that up to 92-94% of human multiexon genes undergo alternative splicing (Chen et al., 2012; Houseley et al., 2009) and mutations in RNA modification enzymes have been associated with over 100 different human diseases (Ketkar et al., 2023). Additionally, the estimated median mRNA half-life in humans is 10 hours, with different functional groups of mRNA decaying at different rates (Yang et al., 2003), such as E. coli RNA with a half-life of only 5-7 minutes.

This characteristic, in addition to the relative abundance and ability to detect isoforms and unexpressed genes, make RNA an attractive basis for molecular tests. The stability of mRNA has also been found to alter gene expression and mRNA life span, and RNA viruses have been found to evade degradation by maintaining mRNA stability (Houseley et al., 2009; Moon et al., 2012; Vasudevan et al., 2007), which further validate our focus on stability as a potential biomarker.

Our results demonstrated that the stability of reads from different genes varied. Because one gene will have a lower stability relative to another from this conclusion, it is possible that one gene was underrepresented in the RNA-Seq analysis. If true, there will be a significant impact on our interpretation of RNA-Seq results.

Genes that may have been regarded as lowly expressed and disregarded to focus on seemingly highly expressed genes may be new targets to be reanalyzed. The contributions of these genes to cellular function may have been underestimated. Additionally, the stability of RNA could impact crucial aspects of a disease’s progression.

Viral load, the severity of the infection, and the effectiveness of current treatments could be influenced by RNA stability, and it remains to be seen if this relationship exists. These studies can be performed for not just SARS Cov-2 infections but for other infections as well. For the purposes of utilizing RNA stability to expand upon RNA-Seq, establishing this relationship between RNA-Seq and RNA stability will require a larger experimental research effort that was not conducted in our study. Such studies may utilize in vitro experimental approaches to assess this potential relationship more directly or the utilization of larger data sets. The basis for such an effort has been established by our study, especially with the validation of free energy evaluation and motif analysis as methods to accomplish further studies.

Within the *N* gene, we found that reads from different regions also differed in stability. Because all these reads came from the *N* gene, differences in the quantity of duplicates did not arise from the expression of the *N* gene itself but the abundance of potentially alternatively spliced RNA and stability of RNA fragments. One of these sequences in the *N* gene was heavily duplicated with 328 repeats. More research should be conducted to elucidate the potential relationships alternative splicing or stability has with duplicated reads in RNA-Seq.

Additionally, our motif analysis demonstrated 7 motifs that corresponded to destabilization of the RNA and a single motif that corresponded to stabilization of the RNA. These motifs were not present in the most duplicated read sequence, but there may still be motifs in that read sequence that were not detected by our analysis. Potentially, the most repeated sequence has motifs that confer increased stability or an increased chance to be detected by RNA-Seq, but it was not discovered because the dataset was not large enough.

There may not have been enough sequences to establish the basis that there was a motif in the sequence, demonstrating the need to gather a larger diverse dataset. Further analysis of RNA motifs may become crucial with follow-up research. Additional analysis with varying mutations of these motifs could be conducted to further evaluate the role these motifs play in read sequence stability.

Our motif analysis utilized both the established motif analysis tool MEME-ChIP as well as the new motif analysis tool XSTREME because MEME-ChIP is optimized for sequences larger than our average sequence length. However, MEME-ChIP discovered 3 of the 8 motifs that affected RNA stability that XSTREME could not. It is worth noting that these two different motif analysis tools discovered different motifs, and both should be utilized for follow-up research projects.

Because our experimental analysis did not uniformly control genetic expression, including sub-genomic RNA expression, or alternative splicing, it remains unclear if the differences in stability observed in our study affected the RNA-Seq results. Also, the low read count and limited sequencing hinders the generality of our conclusions. However, our study with the discovery of new motifs that may potentially confer increased or decreased stability for RNA serves as the foundation for a future potential study that will assess this research question in a controlled experimental setting.

Utilizing the motifs that we have discovered to potentially alter stability while also limiting differing gene expression levels and alternative splicing, the relationship between stability and RNA-Seq read duplications can be further elucidated, allowing for further analysis, and broadening of research implications of the already well-known RNA-Seq experiment.

In conclusion, we have demonstrated that free energy evaluations and motif analysis are promising methods to determine the stability of secondary structure which may be relevant for developing RNA primers for RNA-Seq-informed PCR. If the gene is expressed at a high level and the transcript is stable, this makes for an ideal RNA sequence to be used in a PCR. Reliable RNA primers can facilitate the use of RNA-Seq-informed PCR in a more efficient diagnosis and treatment of sepsis.

## 6 Author Contributions

AL initially drafted and revised this submission. MC provided the covid-19 data. AF, SM, AL and ER helped to perform the RNA-seq and RNA-seq analysis. ER and JS edited the manuscript several times. SM, AF, and ER provided guidance throughout the conception and execution of this project. All authors reviewed this manuscript.

## 8 Acknowledgments

Authors acknowledges support from US National Institutes of Health: P20 GM121344 (AF, GN), R35 GM118097 (AA), R01 GM 127472 (WF), R35 GM 142638 (SM).

## 9 Data Availability Statement

The data presented in this study are deposited in the Brown Digital Repository. Ask Brown University for access to the analyzed data.

## References

1. Bailey, T. L., J. Johnson, C. E. Grant and W. S. Noble (2015). “The MEME Suite.” Nucleic Acids Res 43(W1): W39–49.

2. Byron, S. A., K. R. Van Keuren-Jensen, D. M. Engelthaler, J. D. Carpten and D. W. Craig (2016). “Translating RNA sequencing into clinical diagnostics: opportunities and challenges.” Nat Rev Genet 17(5): 257–271.

3. Camargo, J. F., A. A. Ahmed, M. S. Lindner, M. I. Morris, S. Anjan, A. D. Anderson, C. E. Prado, S. C. Dalai, O. V. Martinez and K. V. Komanduri (2019). “Next-generation sequencing of microbial cell-free DNA for rapid noninvasive diagnosis of infectious diseases in immunocompromised hosts.” F1000Res 8: 1194.

4. Chen, L., J. M. Tovar-Corona and A. O. Urrutia (2012). “Alternative splicing: a potential source of functional innovation in the eukaryotic genome.” Int J Evol Biol 2012: 596274.

5. Covert, K., E. Bashore, M. Edds and P. O. Lewis (2021). “Utility of the respiratory viral panel as an antimicrobial stewardship tool.” J Clin Pharm Ther 46(2): 277–285.

6. Deschamps-Francoeur, G., J. Simoneau and M. S. Scott (2020). “Handling multi-mapped reads in RNA-seq.” Comput Struct Biotechnol J 18: 1569–1576.

7. Ding, Y., C. Y. Chan and C. E. Lawrence (2005). “RNA secondary structure prediction by centroids in a Boltzmann weighted ensemble.” RNA 11(8): 1157–1166.

8. Doshi, K. J., J. J. Cannone, C. W. Cobaugh and R. R. Gutell (2004). “Evaluation of the suitability of free-energy minimization using nearest-neighbor energy parameters for RNA secondary structure prediction.” BMC Bioinformatics 5: 105.

9. Fredericks, A. M., M. S. Jentzsch, W. G. Cioffi, M. Cohen, W. G. Fairbrother, S. J. Gandhi, E. O. Harrington, G. J. Nau, J. S. Reichner, C. E. Ventetuolo, M. M. Levy, A. Ayala and S. F. Monaghan (2022). “Deep RNA sequencing of intensive care unit patients with COVID-19.” Sci Rep 12(1): 15755.

10. Fu, Y., P. H. Wu, T. Beane, P. D. Zamore and Z. Weng (2018). “Elimination of PCR duplicates in RNA-seq and small RNA-seq using unique molecular identifiers.” BMC Genomics 19(1): 531.

11. Grant, E. C. B. L. T. (2021). “XSTREME: comprehensive motif analysis of biological sequence datasets.” BioRxiv

12. Houseley, J. and D. Tollervey (2009). “The many pathways of RNA degradation.” Cell 136(4): 763–776.

13. Huang, W., D. Wang and Y. F. Yao (2021). “Understanding the pathogenesis of infectious diseases by single-cell RNA sequencing.” Microb Cell 8(9): 208–222.

14. Jonkhout, N., J. Tran, M. A. Smith, N. Schonrock, J. S. Mattick and E. M. Novoa (2017). “The RNA modification landscape in human disease.” RNA 23(12): 1754–1769.

15. Ketkar, S., L. C. Burrage and B. Lee (2023). “RNA Sequencing as a Diagnostic Tool.” JAMA 329(1): 85–86.

16. Lorenz, R., S. H. Bernhart, C. Honer Zu Siederdissen, H. Tafer, C. Flamm, P. F. Stadler and I. L. Hofacker (2011). “ViennaRNA Package 2.0.” Algorithms Mol Biol 6: 26.

17. Machanick, P. and T. L. Bailey (2011). “MEME-ChIP: motif analysis of large DNA datasets.” Bioinformatics 27(12): 1696–1697.

18. McCaskill, J. S. (1990). “The equilibrium partition function and base pair binding probabilities for RNA secondary structure.” Biopolymers 29(6-7): 1105–1119.

19. Moon, S. L., M. D. Barnhart and J. Wilusz (2012). “Inhibition and avoidance of mRNA degradation by RNA viruses.” Curr Opin Microbiol 15(4): 500–505.

20. Mortazavi, A., B. A. Williams, K. McCue, L. Schaeffer and B. Wold (2008). “Mapping and quantifying mammalian transcriptomes by RNA-Seq.” Nat Methods 5(7): 621–628.

21. Oliva, G., T. Sahr and C. Buchrieser (2015). “Small RNAs, 5’ UTR elements and RNA-binding proteins in intracellular bacteria: impact on metabolism and virulence.” FEMS Microbiol Rev 39(3): 331–349.

22. Palavecino, E. L. (2020). “Rapid Methods for Detection of MRSA in Clinical Specimens.” Methods Mol Biol 2069: 29–45.

23. Papenfort, K. and J. Vogel (2014). “Small RNA functions in carbon metabolism and virulence of enteric pathogens.” Front Cell Infect Microbiol 4: 91.

24. Peymani, F., A. Farzeen and H. Prokisch (2022). “RNA sequencing role and application in clinical diagnostic.” Pediatr Investig 6(1): 29–35.

25. Trotta, E. (2014). “On the normalization of the minimum free energy of RNAs by sequence length.” PLoS One 9(11): e113380.

26. Vasudevan, S. and J. A. Steitz (2007). “AU-rich-element-mediated upregulation of translation by FXR1 and Argonaute 2.” Cell 128(6): 1105–1118.

27. Wu, W., Y. Cheng, H. Zhou, C. Sun and S. Zhang (2023). “The SARS-CoV-2 nucleocapsid protein: its role in the viral life cycle, structure and functions, and use as a potential target in the development of vaccines and diagnostics.” Virol J 20(1): 6.

28. Wuchty, S., W. Fontana, I. L. Hofacker and P. Schuster (1999). “Complete suboptimal folding of RNA and the stability of secondary structures.” Biopolymers 49(2): 145–165.

29. Yang, E., E. van Nimwegen, M. Zavolan, N. Rajewsky, M. Schroeder, M. Magnasco and J. E. Darnell, Jr. (2003). “Decay rates of human mRNAs: correlation with functional characteristics and sequence attributes.” Genome Res 13(8): 1863–1872.

30. Zhou, T. L., H. X. Chen, Y. Z. Wang, S. J. Wen, P. H. Dao, Y. H. Wang and M. F. Chen (2023). “Single-cell RNA sequencing reveals the immune microenvironment and signaling networks in cystitis glandularis.” Front Immunol 14: 1083598.

31. Zuker, M. and P. Stiegler (1981). “Optimal computer folding of large RNA sequences using thermodynamics and auxiliary information.” Nucleic Acids Res 9(1): 133–148.

